# Assessment of *ANG* variants in Parkinson’s disease

**DOI:** 10.1101/2020.10.26.20191510

**Authors:** Francis P. Grenn, Anni Moore, Sara Bandres-Ciga, Lynne Krohn, Cornelis Blauwendraat, on behalf of the International Parkinson’s Disease Genomics Consortium (IPDGC)

**Author notes:** **Corresponding Author:** Cornelis Blauwendraat,. Porter Neuroscience Center, 35 Convent Drive, Bethesda, MD 20892, USA.

## Abstract

Genetic risk factors are occasionally shared between different neurodegenerative diseases. Previous studies have linked *ANG*, a gene encoding angiogenin, to both Parkinson’s disease (PD) and amyotrophic lateral sclerosis (ALS). Functional studies suggest *ANG* plays a neuroprotective role in both PD and ALS by reducing cell death. We further explored the genetic association between *ANG* and PD by analyzing genotype data from the International Parkinson’s Disease Genomics Consortium (IPDGC) (14,671 cases and 17,667 controls) and whole genome sequencing (WGS) data from the Accelerating Medicines Partnership - Parkinson’s disease initiative (AMP-PD, https://amp-pd.org/) (1,647 cases and 1,050 controls). Our analysis did not replicate the findings of previous studies and found no significant association between *ANG* variants and PD risk.

## Introduction

Parkinson’s disease (PD) is a neurodegenerative disease characterized by loss of dopaminergic neurons in the substantia nigra leading to symptoms of tremor, rigidity and slowed movement, and is the second most common neurodegenerative disease in the world. Both sporadic and familial forms of PD exist, and much work has been done to identify the environmental and genetic risk factors behind this disease. Over 20 genes have been associated with PD or parkinsonism in recent years, and the largest genome wide association studies for PD risk have identified 92 PD risk variants across 80 loci, explaining 16-36% of the heritable risk of PD (Blauwendraat et al., 2020; Foo et al., 2020; Nalls et al., 2019).

It is not uncommon to find genetic variations associated with multiple neurodegenerative disorders, suggesting shared pathways between diseases (Tan et al., 2019). For example, common variations in *MAPT* have been associated with PD (Zabetian et al., 2007), amyotrophic lateral sclerosis (ALS) (Karch et al., 2018) and Alzheimer’s disease (AD) (Ferrari et al., 2017), and variations in *GBA* have been associated with PD (Sidransky et al., 2009) and Gaucher disease (Riboldi and Di Fonzo, 2019). Therefore, the interrogation of genes common to multiple neurodegenerative disorders is a logical next step in the identification of novel PD risk variants.

One such candidate exists in *ANG*, a gene thought to confer a large risk for both ALS and PD (Rayaprolu et al., 2012; van Es et al., 2011). However, studies in Asian populations have suggested there is no link between *ANG* variants and PD (Chen et al., 2014; Liu et al., 2013). *ANG* encodes angiogenin, a small protein that plays a role in the angiogenesis pathway, which forms new blood vessels. Angiogenin and its related pathway are thought to play a role in cancer and placental development (Amankwah et al., 2012; Pavlov et al., 2014). An in vitro study has shown that angiogenin has a neuroprotective effect on motor neurons (Subramanian et al., 2008). ALS associated *ANG* variants are suggested to potentiate neuronal death through inhibition of the PI3K-Akt pathway (Kieran et al., 2008). A PD mouse model has also shown this gene has a neuroprotective effect on dopaminergic neurons (Steidinger et al., 2011). This neuroprotective effect is suggested to be lost when *ANG* is mutated, decreasing the viability of motor neurons (Wu et al., 2007). These findings are of relevance because PD is characterized by the loss of dopaminergic neurons and ALS is characterized by the loss of motor neurons. Interestingly, angiogenin levels have been found to be elevated in the blood serum of ALS patients, but not in PD patients (van Es et al., 2014). This suggests angiogenin may play a larger role elsewhere, such as in the basal ganglia, a brain structure often associated with PD. Structural work has shown ten *ANG* coding variants are associated with a decrease in angiogenin activity, and one coding variant, p.Arg145Cys, is associated with an increase in activity (Bradshaw et al., 2017).

To date, *ANG* variants have not been associated with either ALS or PD through genome wide association studies (GWAS) (Nalls et al., 2019; Nicolas et al., 2018), despite previous studies suggesting *ANG* is associated with risk for these diseases (Rayaprolu et al., 2012; van Es et al., 2011). Here we scrutinize *ANG* variants in two large PD datasets to assess whether *ANG* variants contribute to PD risk in individuals of European ancestry.

## Methods

We mined whole-genome sequencing (WGS) data from the Accelerating Medicines Partnership - Parkinson’s disease initiative (AMP-PD, https://amp-pd.org/) which included 1,647 cases and 1,050 healthy controls from cohorts including the Fox Investigation for New Discovery of Biomarkers (BioFIND), the Parkinson’s Progression Markers Initiative (PPMI), the Harvard Biomarker Study (HBS), and the Parkinson’s Disease Biomarkers Program (PDBP). We also looked at *ANG* variants in genotype data from the International Parkinson’s Disease Genomics Consortium (IPDGC) which included 14,671 cases and 17,667 healthy controls. Variants were annotated from both datasets using ANNOVAR (Wang et al., 2010). Variant frequencies in non-Finnish European populations were obtained from the hg38 gnomAD v3.0 dataset (Karczewski et al., 2020). PLINK 1.9 was used to perform Fisher’s exact test to identify significant variants (Purcell et al., 2007). Rare variant burden tests were performed using RVTESTS (Zhan et al., 2016). We further analysed existing summary statistics including the latest GWAS meta-analyses for PD risk and age of onset (Blauwendraat et al., 2019; Nalls et al., 2019) and additionally assessed public summary statistics from the most recent ALS GWAS (Nicolas et al., 2018).

Variants identified by amino acid change from previous studies including Van Es et al. and Rayaprolu et al. did not initially match any variants identified in AMP-PD data due to differences in nomenclature. To resolve this we mapped each variant amino acid change to the angiogenin protein sequence. This sequence was obtained from Ensembl using the ANG-201 ENST00000336811.10 transcript (Yates et al., 2020). We found that the reported amino acid changes from Van Es et al. and Rayaprolu et al. were offset by 25 or 24 amino acids due to numbering differences used for the signal peptide sequence and we accounted for this in our analysis. The code we used for analysis is available on the IPDGC github (https://github.com/ipdgc/IPDGC-Trainees/blob/master/ANG.md).

## Results

We identified a total of 168 *ANG* variants in the AMP-PD WGS data. Nine of these variants were found to be coding. Two of these were synonymous and the other seven were nonsynonymous. The top variant (p=0.017) after performing Fisher’s exact test was not significant after Bonferroni correction for multiple tests (p=0.05/168=2.97E-4) (Supplementary Table 1).

We compared the nine identified *ANG* coding variants to variants from two other studies (Rayaprolu et al., 2012; van Es et al., 2011) (Supplementary Table 2). All nonsynonymous variants were rare (MAF<0.01). Allele frequencies did not differ significantly from gnomAD non-Finnish European allele frequencies although most variants were too rare to reliably test individually.

After excluding two synonymous variants, rs11701 (p.G110=) and rs2228653 (p.T121=), from these nine, we observed a frequency of 0.39% in PD cases and 0.48% in controls. Van Es et al. also removed two common variants, rs121909536 (K41I) and rs121909541 (p.I70V), from their analysis. After removing these variants from our data the frequencies were 0.15% in PD cases and 0.19% in controls. This is in contrast with the previously found 0.45% in PD cases and 0.04% in controls (van Es et al., 2011).

Burden tests using variants with minor allele frequency less than 0.03 gave no significant results when using all variants (N variants=72; CMC p=0.493, Fp p=0.509, MB p=0.880, Skat p=0.454, SkatO p=0.523, Zeggini p=0.395). Likewise, there were no significant results when doing the same test on only coding variants (N variants=9; CMC p=0.866, Fp p=0.510, MB p=0.820, Skat p=0.436, SkatO p=0.556, Zeggini p=0.868).

Twenty-six *ANG* variants were found using the IPDGC imputed genotype data, all of which were non-coding (Supplementary Table 1). No significant association between *ANG* variants and PD risk (Figure 1A) or onset (Supplementary Figure 1) was found in data from the latest PD risk GWAS or in the PD age of onset GWAS (Blauwendraat et al., 2019; Nalls et al., 2019). No variants had a minor allele frequency less than 0.03, so the threshold was increased to 0.05 for burden tests. Only two variants were included at this threshold, which also gave no significant results (N=2; CMC p=0.893, Fp p=0.960, MB p=0.948, Skat p=0.980, SkatO p=1, Zeggini p=0.842). Additionally, no GWAS signal of interest is identified in the most recent ALS GWAS (Figure 1B) (Nicolas et al., 2018).

**Figure 1.**
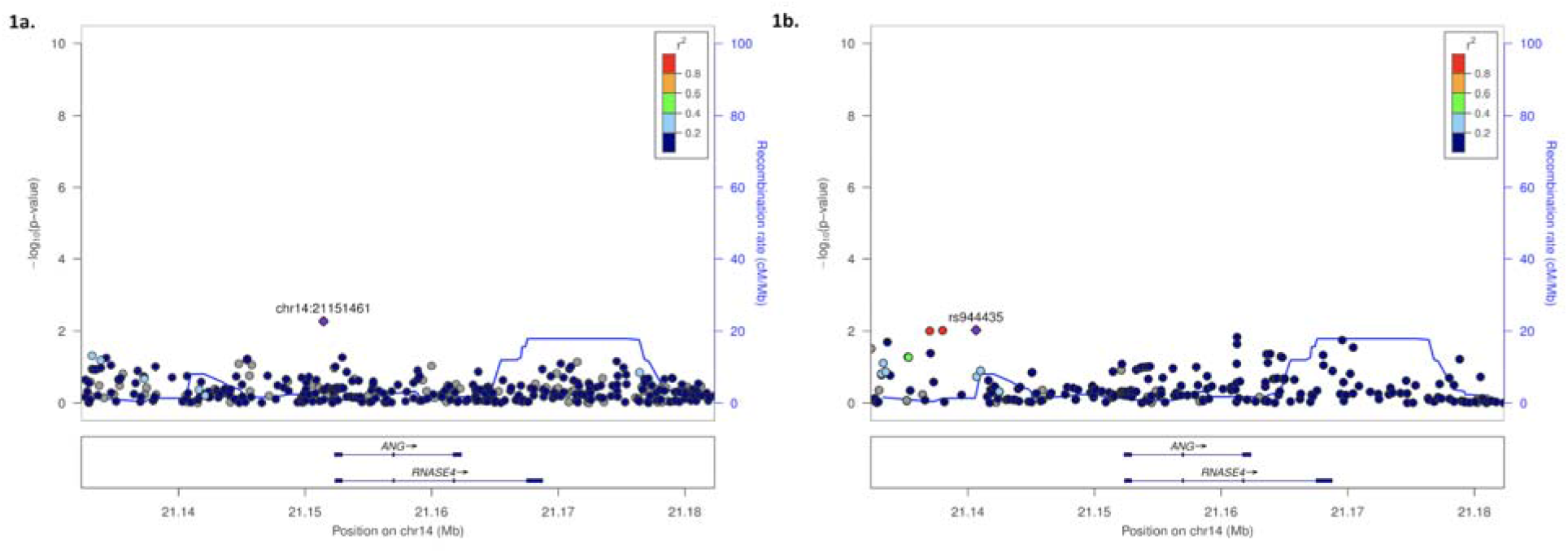
Locus zoom plots for *ANG* and PD risk and ALS risk. The -log_10_(p-value) of variants on or near *ANG* are shown on the y-axis, and base-pair position of each variant is on the x-axis. P-values are taken from the PD risk GWAS (Figure 1A) and the ALS risk GWAS (Figure 1B). Variants are colored by their R^2^ linkage disequilibrium color with respect to the variant with the lowest p-value on this plot. Recombination rates are included in blue (Pruim et al., 2010).

## Discussion

Rare coding variants in *ANG* have been reported to be associated with PD (van Es et al., 2011). Here, our goal was to further explore the role of *ANG* in PD by analyzing large datasets from IPDGC and AMP-PD. Our study shows no significant enrichment of *ANG* single variants in PD cases or controls in either of these datasets. Rare variant burden tests also gave no significant results for *ANG*. Our analysis provides no evidence to support the hypothesis that genetic variation of *ANG* plays a role in PD risk or age at onset.

The nine coding *ANG* variants we identified were from AMP-PD WGS data. This dataset included fewer samples (1,647 PD; 1,050 controls) than the Van Es et al. study (6,471 ALS;3,146 PD;7,668 controls) which identified a total of 29 unique *ANG* coding variants. However, the frequency of *ANG* coding variants detected in the AMP-PD data is 0.15% in PD cases and 0.19% in controls which is different from the previously found 0.45% in PD cases and 0.04% in controls (van Es et al., 2011). Using the Genetic Association Study Power Calculator, we calculated a genotype relative risk of 3.7 at a statistical power of 0.8 (Supplementary Figure 2) (Johnson and Abecasis, n.d.). The genotype relative risk increased to 6.7 when using a statistical power of 0.95. This is comparable to the PD odds ratio of 6.7 from previous studies, suggesting we have the statistical power required to replicate these findings (van Es et al., 2011). However, the cumulative frequency of *ANG* variants identified in AMP-PD data was not significantly different as previously reported. A larger sample size may be needed to identify the missing coding variants so the role of *ANG* in PD can be assessed on an even larger scale.

Overall, despite some potentially interesting functional experiments supporting the neuroprotective effect of angiogenin, we cannot replicate the genetic association between *ANG* coding variants and PD. Therefore, we cannot conclude that *ANG* variants play a role in PD, which is in line with previous studies done in Asian populations (Chen et al., 2014; Liu et al., 2013).

## Data Availability

data is provided in supplementary tables and code used to generate the provided data is on our github (https://github.com/ipdgc/IPDGC-Trainees/blob/master/ANG.md).

## Acknowledgements

We would like to thank all of the subjects who donated their time and biological samples to be a part of this study. We also would like to thank all members of the International Parkinson’s Disease Genomics Consortium (IPDGC). For a complete overview of members, acknowledgements and funding, please see http://pdgenetics.org/partners. This work was supported in part by the Intramural Research Programs of the National Institute of Neurological Disorders and Stroke (NINDS), the National Institute on Aging (NIA), and the National Institute of Environmental Health Sciences (NIEHS), all part of the National Institutes of Health, Department of Health and Human Services; project numbers 1ZIA-NS003154, Z01-AG000949-02 and Z01-ES101986. In addition, this work was supported by the Department of Defense (award W81XWH-09-2-0128), and The Michael J Fox Foundation for Parkinson’s Research. Data used in the preparation of this article were obtained from the AMP PD Knowledge Platform. For up-to-date information on the study, visit https://www.amp-pd.org. AMP PD – a public-private partnership – is managed by the FNIH and funded by Celgene, GSK, the Michael J. Fox Foundation for Parkinson’s Research, the National Institute of Neurological Disorders and Stroke, Pfizer, and Verily. We would like to thank AMP-PD for the publicly available whole-genome sequencing data, including cohorts from the Fox Investigation for New Discovery of Biomarkers (BioFIND), the Parkinson’s Progression Markers Initiative (PPMI), and the Parkinson’s Disease Biomarkers Program (PDBP). The Parkinson’s Disease Biomarker Program (PDBP) consortium is supported by the National Institute of Neurological Disorders and Stroke (NINDS) at the National Institutes of Health. A full list of PDBP investigators can be found at https://pdbp.ninds.nih.gov/policy. Harvard Biomarker Study (HBS) is a collaboration of HBS investigators (full list of HBS investigators found at https://www.bwhparkinsoncenter.org/biobank) and funded through philanthropy and NIH and Non-NIH funding sources. The HBS Investigators have not participated in reviewing the data analysis or content of the manuscript. This work utilized the computational resources of the NIH HPC Biowulf cluster (http://hpc.nih.gov).

## References

Amankwah, E.K., Sellers, T.A., Park, J.Y., 2012. Gene variants in the angiogenesis pathway and prostate cancer. Carcinogenesis 33, 1259–1269.

Blauwendraat, C., Heilbron, K., Vallerga, C.L., Bandres-Ciga, S., von Coelln, R., Pihlstrøm, L., Simón-Sánchez, J., Schulte, C., Sharma, M., Krohn, L., Siitonen, A., Iwaki, H., Leonard, H., Noyce, A.J., Tan, M., Gibbs, J.R., Hernandez, D.G., Scholz, S.W., Jankovic, J., Shulman, L.M., Lesage, S., Corvol, J.-C., Brice, A., van Hilten, J.J., Marinus, J., 23andMe Research Team, Eerola-Rautio, J., Tienari, P., Majamaa, K., Toft, M., Grosset, D.G., Gasser, T., Heutink, P., Shulman, J.M., Wood, N., Hardy, J., Morris, H.R., Hinds, D.A., Gratten, J., Visscher, P.M., Gan-Or, Z., Nalls, M.A., Singleton, A.B., International Parkinson’s Disease Genomics Consortium (IPDGC), 2019. Parkinson’s disease age at onset genome-wide association study: Defining heritability, genetic loci, and α-synuclein mechanisms. Mov. Disord. 34, 866–875.

Blauwendraat, C., Nalls, M.A., Singleton, A.B., 2020. The genetic architecture of Parkinson’s disease. Lancet Neurol. 19, 170–178.

Bradshaw, W.J., Rehman, S., Pham, T.T.K., Thiyagarajan, N., Lee, R.L., Subramanian, V., Ravi Acharya, K., 2017. Structural insights into human angiogenin variants implicated in Parkinson’s disease and Amyotrophic Lateral Sclerosis. Scientific Reports. https://doi.org/10.1038/srep41996

Chen, M.-L., Wu, R.-M., Tai, C.-H., Lin, C.-H., 2014. Mutational analysis of angiogenin gene in Parkinson’s disease. PLoS One 9, e112661.

Ferrari, R., Wang, Y., Vandrovcova, J., Guelfi, S., Witeolar, A., Karch, C.M., Schork, A.J., Fan, C.C., Brewer, J.B., International FTD-Genomics Consortium (IFGC), International Parkinson’s Disease Genomics Consortium (IPDGC), International Genomics of Alzheimer’s Project (IGAP), Momeni, P., Schellenberg, G.D., Dillon, W.P., Sugrue, L.P., Hess, C.P., Yokoyama, J.S., Bonham, L.W., Rabinovici, G.D., Miller, B.L., Andreassen, O.A., Dale, A.M., Hardy, J., Desikan, R.S., 2017. Genetic architecture of sporadic frontotemporal dementia and overlap with Alzheimer’s and Parkinson’s diseases. J. Neurol. Neurosurg. Psychiatry 88, 152–164.

Foo, J.N., Chew, E.G.Y., Chung, S.J., Peng, R., Blauwendraat, C., Nalls, M.A., Mok, K.Y., Satake, W., Toda, T., Chao, Y., Tan, L.C.S., Tandiono, M., Lian, M.M., Ng, E.Y., Prakash, K.-M., Au, W.-L., Meah, W.-Y., Mok, S.Q., Annuar, A.A., Chan, A.Y.Y., Chen, L., Chen, Y., Jeon, B.S., Jiang, L., Lim, J.L., Lin, J.-J., Liu, C., Mao, C., Mok, V., Pei, Z., Shang, H.-F., Shi, C.-H., Song, K., Tan, A.H., Wu, Y.-R., Xu, Y.-M., Xu, R., Yan, Y., Yang, J., Zhang, B., Koh, W.-P., Lim, S.-Y., Khor, C.C., Liu, J., Tan, E.-K., 2020. Identification of Risk Loci for Parkinson Disease in Asians and Comparison of Risk Between Asians and Europeans: A Genome-Wide Association Study. JAMA Neurol. https://doi.org/10.1001/jamaneurol.2020.0428

Johnson, J.L., Abecasis, G.R., n.d. GAS Power Calculator: web-based power calculator for genetic association studies. https://doi.org/10.1101/164343

Karch, C.M., Wen, N., Fan, C.C., Yokoyama, J.S., Kouri, N., Ross, O.A., Höglinger, G., Müller, U., Ferrari, R., Hardy, J., Schellenberg, G.D., Sleiman, P.M., Momeni, P., Hess, C.P., Miller, B.L., Sharma, M., Van Deerlin, V., Smeland, O.B., Andreassen, O.A., Dale, A.M., Desikan, R.S., International Frontotemporal Dementia (FTD)–Genomics Consortium, International Collaboration for Frontotemporal Dementia, Progressive Supranuclear Palsy (PSP) Genetics Consortium, and International Parkinson’s Disease Genomics Consortium, 2018. Selective Genetic Overlap Between Amyotrophic Lateral Sclerosis and Diseases of the Frontotemporal Dementia Spectrum. JAMA Neurol. 75, 860–875.

Karczewski, K.J., Francioli, L.C., Tiao, G., Cummings, B.B., Alföldi, J., Wang, Q., Collins, R.L., Laricchia, K.M., Ganna, A., Birnbaum, D.P., Gauthier, L.D., Brand, H., Solomonson, M., Watts, N.A., Rhodes, D., Singer-Berk, M., England, E.M., Seaby, E.G., Kosmicki, J.A., Walters, R.K., Tashman, K., Farjoun, Y., Banks, E., Poterba, T., Wang, A., Seed, C., Whiffin, N., Chong, J.X., Samocha, K.E., Pierce-Hoffman, E., Zappala, Z., O’Donnell-Luria, A.H., Minikel, E.V., Weisburd, B., Lek, M., Ware, J.S., Vittal, C., Armean, I.M., Bergelson, L., Cibulskis, K., Connolly, K.M., Covarrubias, M., Donnelly, S., Ferriera, S., Gabriel, S., Gentry, J., Gupta, N., Jeandet, T., Kaplan, D., Llanwarne, C., Munshi, R., Novod, S., Petrillo, N., Roazen, D., Ruano-Rubio, V., Saltzman, A., Schleicher, M., Soto, J., Tibbetts, K., Tolonen, C., Wade, G., Talkowski, M.E., Genome Aggregation Database Consortium, Neale, B.M., Daly, M.J., MacArthur, D.G., 2020. The mutational constraint spectrum quantified from variation in 141,456 humans. Nature 581, 434–443.

Kieran, D., Sebastia, J., Greenway, M.J., King, M.A., Connaughton, D., Concannon, C.G., Fenner, B., Hardiman, O., Prehn, J.H.M., 2008. Control of motoneuron survival by angiogenin. J. Neurosci. 28, 14056–14061.

Liu, B., Zhang, Y., Wang, Y., Xiao, Q., Yang, Q., Wang, G., Ma, J., Zhao, J., Quinn, T.J., Chen, S.-D., Liu, J., 2013. Angiogenin variants are not associated with Parkinson’s disease in the ethnic Chinese population. Parkinsonism & Related Disorders. https://doi.org/10.1016/j.parkreldis.2012.11.016

Nalls, M.A., Blauwendraat, C., Vallerga, C.L., Heilbron, K., Bandres-Ciga, S., Chang, D., Tan, M., Kia, D.A., Noyce, A.J., Xue, A., Bras, J., Young, E., von Coelln, R., Simón-Sánchez, J., Schulte, C., Sharma, M., Krohn, L., Pihlstrøm, L., Siitonen, A., Iwaki, H., Leonard, H., Faghri, F., Gibbs, J.R., Hernandez, D.G., Scholz, S.W., Botia, J.A., Martinez, M., Corvol, J.-C., Lesage, S., Jankovic, J., Shulman, L.M., Sutherland, M., Tienari, P., Majamaa, K., Toft, M., Andreassen, O.A., Bangale, T., Brice, A., Yang, J., Gan-Or, Z., Gasser, T., Heutink, P., Shulman, J.M., Wood, N.W., Hinds, D.A., Hardy, J.A., Morris, H.R., Gratten, J., Visscher, P.M., Graham, R.R., Singleton, A.B., 23andMe Research Team, System Genomics of Parkinson’s Disease Consortium, International Parkinson’s Disease Genomics Consortium, 2019. Identification of novel risk loci, causal insights, and heritable risk for Parkinson’s disease: a meta-analysis of genome-wide association studies. Lancet Neurol. 18, 1091–1102.

Nicolas, A., Kenna, K.P., Renton, A.E., Ticozzi, N., Faghri, F., Chia, R., Dominov, J.A., Kenna, B.J., Nalls, M.A., Keagle, P., Rivera, A.M., van Rheenen, W., Murphy, N.A., van Vugt, J.J.F.A., Geiger, J.T., Van der Spek, R.A., Pliner, H.A., Shankaracharya, Smith, B.N., Marangi, G., Topp, S.D., Abramzon, Y., Gkazi, A.S., Eicher, J.D., Kenna, A., ITALSGEN Consortium, Mora, G., Calvo, A., Mazzini, L., Riva, N., Mandrioli, J., Caponnetto, C., Battistini, S., Volanti, P., La Bella, V., Conforti, F.L., Borghero, G., Messina, S., Simone, I.L., Trojsi, F., Salvi, F., Logullo, F.O., D’Alfonso, S., Corrado, L., Capasso, M., Ferrucci, L., Genomic Translation for ALS Care (GTAC) Consortium, Moreno, C. de A.M., Kamalakaran, S., Goldstein, D.B., ALS Sequencing Consortium, Gitler, A.D., Harris, T., Myers, R.M., NYGC ALS Consortium, Phatnani, H., Musunuri, R.L., Evani, U.S., Abhyankar, A., Zody, M.C., Answer ALS Foundation, Kaye, J., Finkbeiner, S., Wyman, S.K., LeNail, A., Lima, L., Fraenkel, E., Svendsen, C.N., Thompson, L.M., Van Eyk, J.E., Berry, J.D., Miller, T.M., Kolb, S.J., Cudkowicz, M., Baxi, E., Clinical Research in ALS and Related Disorders for Therapeutic Development (CReATe) Consortium, Benatar, M., Taylor, J.P., Rampersaud, E., Wu, G., Wuu, J., SLAGEN Consortium, Lauria, G., Verde, F., Fogh, I., Tiloca, C., Comi, G.P., Sorarù, G., Cereda, C., French ALS Consortium, Corcia, P., Laaksovirta, H., Myllykangas, L., Jansson, L., Valori, M., Ealing, J., Hamdalla, H., Rollinson, S., Pickering-Brown, S., Orrell, R.W., Sidle, K.C., Malaspina, A., Hardy, J., Singleton, A.B., Johnson, J.O., Arepalli, S., Sapp, P.C., McKenna-Yasek, D., Polak, M., Asress, S., Al-Sarraj, S., King, A., Troakes, C., Vance, C., de Belleroche, J., Baas, F., Ten Asbroek, A.L.M.A., Muñoz-Blanco, J.L., Hernandez, D.G., Ding, J., Gibbs, J.R., Scholz, S.W., Floeter, M.K., Campbell, R.H., Landi, F., Bowser, R., Pulst, S.M., Ravits, J.M., MacGowan, D.J.L., Kirby, J., Pioro, E.P., Pamphlett, R., Broach, J., Gerhard, G., Dunckley, T.L., Brady, C.B., Kowall, N.W., Troncoso, J.C., Le Ber, I., Mouzat, K., Lumbroso, S., Heiman-Patterson, T.D., Kamel, F., Van Den Bosch, L., Baloh, R.H., Strom, T.M., Meitinger, T., Shatunov, A., Van Eijk, K.R., de Carvalho, M., Kooyman, M., Middelkoop, B., Moisse, M., McLaughlin, R.L., Van Es, M.A., Weber, M., Boylan, K.B., Van Blitterswijk, M., Rademakers, R., Morrison, K.E., Basak, A.N., Mora, J.S., Drory, V.E., Shaw, P.J., Turner, M.R., Talbot, K., Hardiman, O., Williams, K.L., Fifita, J.A., Nicholson, G.A., Blair, I.P., Rouleau, G.A., Esteban-Pérez, J., García-Redondo, A., Al-Chalabi, A., Project MinE ALS Sequencing Consortium, Rogaeva, E., Zinman, L., Ostrow, L.W., Maragakis, N.J., Rothstein, J.D., Simmons, Z., Cooper-Knock, J., Brice, A., Goutman, S.A., Feldman, E.L., Gibson, S.B., Taroni, F., Ratti, A., Gellera, C., Van Damme, P., Robberecht, W., Fratta, P., Sabatelli, M., Lunetta, C., Ludolph, A.C., Andersen, P.M., Weishaupt, J.H., Camu, W., Trojanowski, J.Q., Van Deerlin, V.M., Brown, R.H., Jr, van den Berg, L.H., Veldink, J.H., Harms, M.B., Glass, J.D., Stone, D.J., Tienari, P., Silani, V., Chiò, A., Shaw, C.E., Traynor, B.J., Landers, J.E., 2018. Genome-wide Analyses Identify KIF5A as a Novel ALS Gene. Neuron 97, 1268–1283.e6.

Pavlov, N., Frendo, J.-L., Guibourdenche, J., Degrelle, S.A., Evain-Brion, D., Badet, J., 2014. Angiogenin expression during early human placental development; association with blood vessel formation. Biomed Res. Int. 2014, 781632.

Pruim, R.J., Welch, R.P., Sanna, S., Teslovich, T.M., Chines, P.S., Gliedt, T.P., Boehnke, M., Abecasis, G.R., Willer, C.J., 2010. LocusZoom: regional visualization of genome-wide association scan results. Bioinformatics 26, 2336–2337.

Purcell, S., Neale, B., Todd-Brown, K., Thomas, L., Ferreira, M.A.R., Bender, D., Maller, J., Sklar, P., de Bakker, P.I.W., Daly, M.J., Sham, P.C., 2007. PLINK: A Tool Set for Whole-Genome Association and Population-Based Linkage Analyses. The American Journal of Human Genetics. https://doi.org/10.1086/519795

Rayaprolu, S., Soto-Ortolaza, A., Rademakers, R., Uitti, R.J., Wszolek, Z.K., Ross, O.A., 2012. Angiogenin variation and Parkinson disease. Ann. Neurol.

Riboldi, G.M., Di Fonzo, A.B., 2019. GBA, Gaucher Disease, and Parkinson’s Disease: From Genetic to Clinic to New Therapeutic Approaches. Cells. https://doi.org/10.3390/cells8040364

Sidransky, E., Nalls, M.A., Aasly, J.O., Aharon-Peretz, J., Annesi, G., Barbosa, E.R., Bar-Shira, A., Berg, D., Bras, J., Brice, A., Chen, C.-M., Clark, L.N., Condroyer, C., De Marco, E.V., Dürr, A., Eblan, M.J., Fahn, S., Farrer, M.J., Fung, H.-C., Gan-Or, Z., Gasser, T., Gershoni-Baruch, R., Giladi, N., Griffith, A., Gurevich, T., Januario, C., Kropp, P., Lang, A.E., Lee-Chen, G.-J., Lesage, S., Marder, K., Mata, I.F., Mirelman, A., Mitsui, J., Mizuta, I., Nicoletti, G., Oliveira, C., Ottman, R., Orr-Urtreger, A., Pereira, L.V., Quattrone, A., Rogaeva, E., Rolfs, A., Rosenbaum, H., Rozenberg, R., Samii, A., Samaddar, T., Schulte, C., Sharma, M., Singleton, A., Spitz, M., Tan, E.-K., Tayebi, N., Toda, T., Troiano, A.R., Tsuji, S., Wittstock, M., Wolfsberg, T.G., Wu, Y.-R., Zabetian, C.P., Zhao, Y., Ziegler, S.G., 2009. Multicenter analysis of glucocerebrosidase mutations in Parkinson’s disease. N. Engl. J. Med. 361, 1651–1661.

Steidinger, T.U., Standaert, D.G., Yacoubian, T.A., 2011. A neuroprotective role for angiogenin in models of Parkinson’s disease. Journal of Neurochemistry. https://doi.org/10.1111/j.1471-4159.2010.07112.x

Subramanian, V., Crabtree, B., Acharya, K.R., 2008. Human angiogenin is a neuroprotective factor and amyotrophic lateral sclerosis associated angiogenin variants affect neurite extension/pathfinding and survival of motor neurons. Hum. Mol. Genet. 17, 130–149.

Tan, S.H., Karri, V., Tay, N.W.R., Chang, K.H., Ah, H.Y., Ng, P.Q., Ho, H.S., Keh, H.W., Candasamy, M., 2019. Emerging pathways to neurodegeneration: Dissecting the critical molecular mechanisms in Alzheimer’s disease, Parkinson’s disease. Biomedicine & Pharmacotherapy. https://doi.org/10.1016/j.biopha.2018.12.101

van Es, M.A., Schelhaas, H.J., van Vught, P.W.J., Ticozzi, N., Andersen, P.M., Groen, E.J.N., Schulte, C., Blauw, H.M., Koppers, M., Diekstra, F.P., Fumoto, K., LeClerc, A.L., Keagle, P., Bloem, B.R., Scheffer, H., van Nuenen, B.F.L., van Blitterswijk, M., van Rheenen, W., Wills, A.-M., Lowe, P.P., Hu, G.-F., Yu, W., Kishikawa, H., Wu, D., Folkerth, R.D., Mariani, C., Goldwurm, S., Pezzoli, G., Van Damme, P., Lemmens, R., Dahlberg, C., Birve, A., Fernández-Santiago, R., Waibel, S., Klein, C., Weber, M., van der Kooi, A.J., de Visser, M., Verbaan, D., van Hilten, J.J., Heutink, P., Hennekam, E.A.M., Cuppen, E., Berg, D., Brown, R.H., Jr, Silani, V., Gasser, T., Ludolph, A.C., Robberecht, W., Ophoff, R.A., Veldink, J.H., Pasterkamp, R.J., de Bakker, P.I.W., Landers, J.E., van de Warrenburg, B.P., van den Berg, L.H., 2011. Angiogenin variants in Parkinson disease and amyotrophic lateral sclerosis. Ann. Neurol. 70, 964–973.

van Es, M.A., Veldink, J.H., Schelhaas, H.J., Bloem, B.R., Sodaar, P., van Nuenen, B.F.L., Verbeek, M., van de Warrenburg, B.P., van den Berg, L.H., 2014. Serum angiogenin levels are elevated in ALS, but not Parkinson’s disease: Table 1. Journal of Neurology, Neurosurgery & Psychiatry. https://doi.org/10.1136/jnnp-2013-307168

Wang, K., Li, M., Hakonarson, H., 2010. ANNOVAR: functional annotation of genetic variants from high-throughput sequencing data. Nucleic Acids Res. 38, e164.

Wu, D., Yu, W., Kishikawa, H., Folkerth, R.D., Iafrate, A.J., Shen, Y., Xin, W., Sims, K., Hu, G.-F., 2007. Angiogenin loss-of-function mutations in amyotrophic lateral sclerosis. Ann. Neurol. 62, 609–617.

Yates, A.D., Achuthan, P., Akanni, W., Allen, J., Allen, J., Alvarez-Jarreta, J., Amode, M.R., Armean, I.M., Azov, A.G., Bennett, R., Bhai, J., Billis, K., Boddu, S., Marugán, J.C., Cummins, C., Davidson, C., Dodiya, K., Fatima, R., Gall, A., Giron, C.G., Gil, L., Grego, T., Haggerty, L., Haskell, E., Hourlier, T., Izuogu, O.G., Janacek, S.H., Juettemann, T., Kay, M., Lavidas, I., Le, T., Lemos, D., Martinez, J.G., Maurel, T., McDowall, M., McMahon, A., Mohanan, S., Moore, B., Nuhn, M., Oheh, D.N., Parker, A., Parton, A., Patricio, M., Sakthivel, M.P., Abdul Salam, A.I., Schmitt, B.M., Schuilenburg, H., Sheppard, D., Sycheva, M., Szuba, M., Taylor, K., Thormann, A., Threadgold, G., Vullo, A., Walts, B., Winterbottom, A., Zadissa, A., Chakiachvili, M., Flint, B., Frankish, A., Hunt, S.E., IIsley, G., Kostadima, M., Langridge, N., Loveland, J.E., Martin, F.J., Morales, J., Mudge, J.M., Muffato, M., Perry, E., Ruffier, M., Trevanion, S.J., Cunningham, F., Howe, K.L., Zerbino, D.R., Flicek, P., 2020. Ensembl 2020. Nucleic Acids Res. 48, D682–D688.

Zabetian, C.P., Hutter, C.M., Factor, S.A., Nutt, J.G., Higgins, D.S., Griffith, A., Roberts, J.W., Leis, B.C., Kay, D.M., Yearout, D., Montimurro, J.S., Edwards, K.L., Samii, A., Payami, H., 2007. Association Analysis of MAPT H1 Haplotype and Subhaplotypes in Parkinson’s Disease. Ann. Neurol. 62. https://doi.org/10.1002/ana.21157

Zhan, X., Hu, Y., Li, B., Abecasis, G.R., Liu, D.J., 2016. RVTESTS: an efficient and comprehensive tool for rare variant association analysis using sequence data. Bioinformatics 32, 1423–1426.

