## Supplementary Figures for "Assessment of *ANG* variants in Parkinson’s disease"

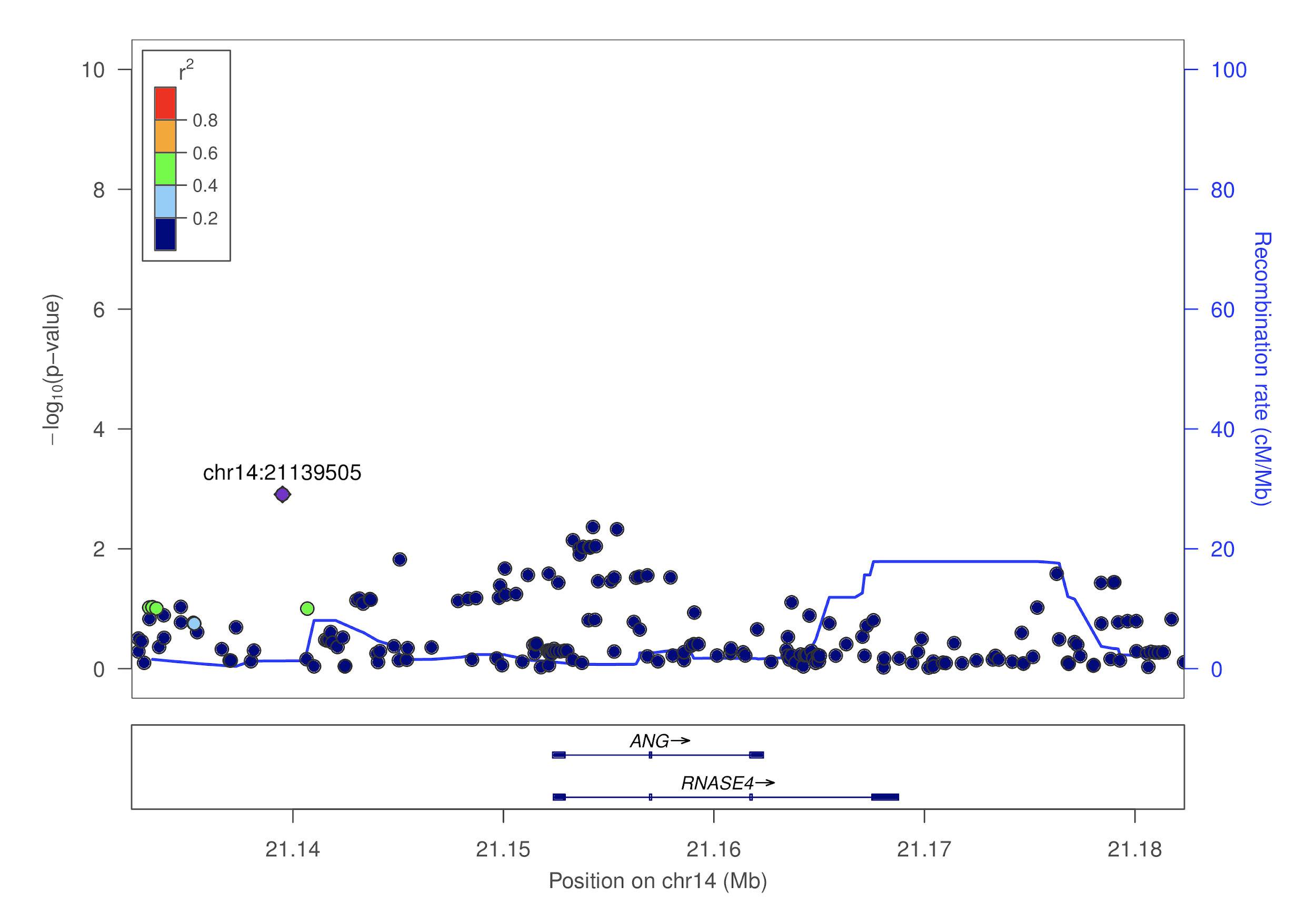

**Supplementary Figure 1. Locus zoom for *ANG* and PD age of onset.** The -log_10_(p-value) of variants on or near *ANG* are shown on the y-axis, and basepair position of each variant is on the x-axis. P-values are taken from the PD age of onset GWAS. Variants are colored by their r^2^ linkage disequilibrium color with respect to the variant with the lowest p-value on this plot. Recombination rates are included in blue.

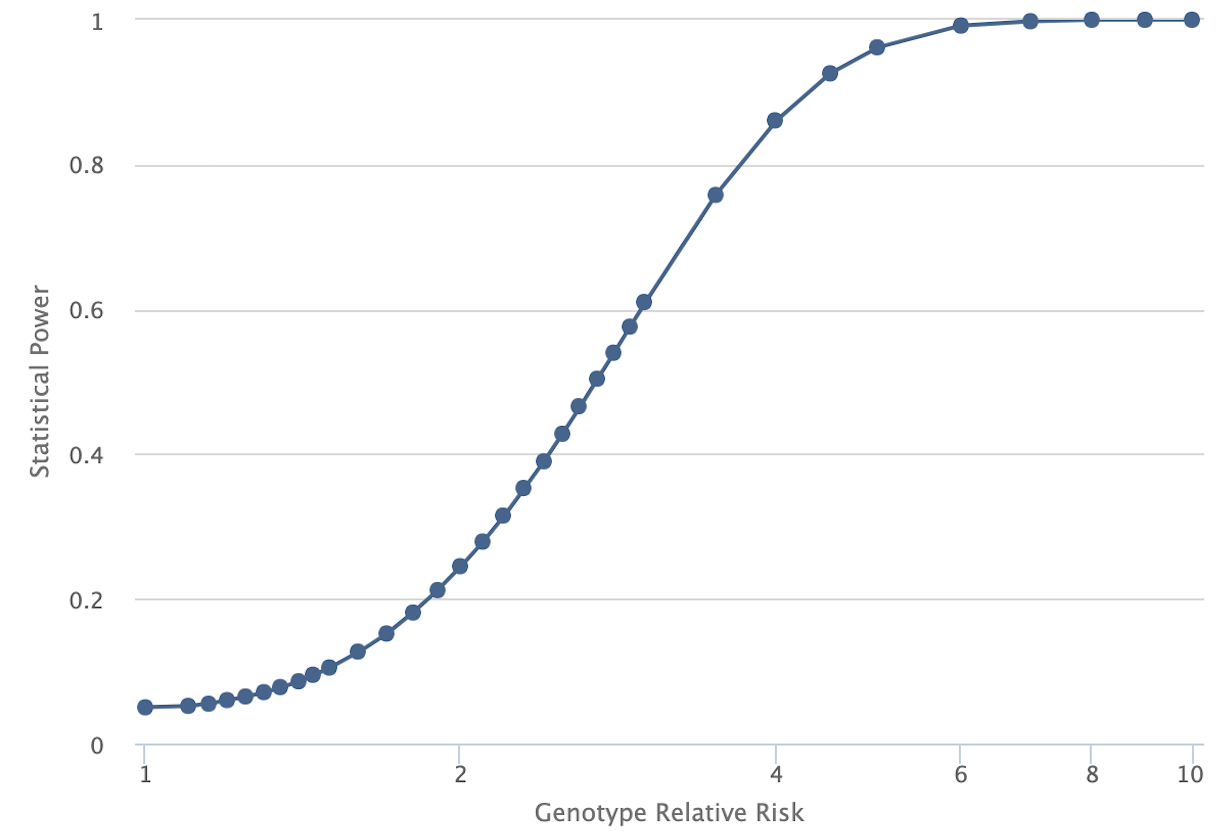

**Supplementary Figure 2. Statistical power versus genotype relative risk in AMP-PD data.** Plot generated from the Genetic Association Study Power Calculator (<http://csg.sph.umich.edu/abecasis/cats/gas_power_calculator/index.html>). Samples sizes were taken from AMP-PD data (1,647 cases, 1,050 controls). A significance level of 0.05 was used with a multiplicative disease model and disease prevalence of 0.01. Disease allele frequency was calculated as 0.0017 (combining cases and controls of AMP-PD data).
